# Detecting recent concussion with multidimensional diffusion imaging: enhanced sensitivity over fractional anisotropy

**DOI:** 10.1101/2025.07.08.25331051

**Authors:** Elizabeth J. Rizor, Neil M. Dundon, Margaret Hayes, Kiana Sabugo, Scott T. Grafton

## Abstract

Despite its classification as a “mild” brain injury, concussion causes a cascade of potentially damaging neural effects. While not often visually detectable on clinical MRI scans, concussion-induced white matter (WM) injury can be detected by diffusion imaging using fractional anisotropy (FA). Though FA is interpreted as a marker of “WM integrity”, it cannot distinguish WM microstructural changes from fiber directionality variation. This limitation highlights the need for alternative imaging biomarkers. Multidimensional Diffusion (MDM) imaging provides estimation of “micro” FA (µFA), which is more robust to fiber directionality variation. Yet, it is unknown whether µFA and other MDM parameters can reliably detect concussion-related injury. We employed MDM imaging to estimate diffusion markers across WM regions in Concussion (N=30, mean age=21.1, sex=13M/17F, scanned within 5 days of injury) and Control (N=32, mean age=22.8, sex=18M/14F) groups. We assessed: 1) whether regional MDM parameter values are credibly associated with concussion, and 2) whether MDM parameter group discriminability outperforms F A. Lower MDM anisotropy (µFA, D^2^_anison_) and greater MDM isotropy (D_iso_, V_ison_; equivalents to mean diffusivity [MD] and MD variance) values were credibly associated with concussion across several regions (94% HDI for β excluded 0). These trends were not unique to areas with crossing fibers. µFA and D^2^_anison_ displayed significantly greater discriminability (higher AUC) when compared to FA (p<0.001). MDM parameters, particularly µFA and D^2^_anison_, are promising biomarkers of concussion-induced WM injury. The lack of specificity to crossing fiber areas challenges the idea that axonal shearing drives concussion-induced injury.

## 1. Introduction

Concussion afflicts millions each year, often sparking a “neurometabolic cascade” of damaging effects, such as white matter (axonal) injury, ionic flux, cell death, and energy disruption (Giza & Hovda, 2014; University of Michigan Health, n.d.). These effects are posited to cause a wide array of physical, cognitive, and psychiatric symptoms that typically peak in the acute phase and subside in weeks or months (Cole & Bailie, 2016).

Despite these significant effects, concussion-induced white matter (WM) injury is typically not visually detectable on computer tomography or clinical MRI scans (Lunkova et al., 2021). However, long-standing evidence has shown that, in patients with concussion, WM abnormalities are detectable via diffusion MRI imaging techniques, which measure the motion of water molecules constrained by WM tracts (Henry et al., 2011; Hulkower et al., 2013; Niogi et al., 2008). Techniques most commonly employed include diffusion tensor imaging to estimate voxel-wise fractional anisotropy (FA) or mean diffusivity (MD); yet, findings with these measures in patients with semi-acute concussion have been contradictory across studies (Dodd et al., 2014). This could be due in part to a threshold below which head impact does not influence diffusion. However, it could also be due to methodological limitations with conventional diffusion imaging.

FA has limited ability to distinguish potential microstructural changes from normal regional variation of fiber orientations and crossings (Jeurissen et al., 2013; Volz et al., 2018). This is particularly important because multidirectional voxels (i.e., voxels with crossing, kissing, and/or fanning fibers) make up more than half of the WM, including segments of tracts traditionally considered to consist mainly of unidirectional/parallel fiber bundles (e.g., corticospinal tract, corpus callosum; Volz et al., 2018). Additionally, there exists long-standing evidence of a propensity for shearing-type injury in crossing fiber areas after traumatic brain injury (Rutgers et al., 2008; Strich, 1961). Given the limitations of FA, there is a need for diffusion imaging biomarkers of concussion that reliably detect (sub-)acute WM injury in tissue of varying fiber directionalities. Such a biomarker could serve as a useful tool to study post-injury therapeutic interventions.

To this end, we employed multidimensional diffusion (MDM) imaging with gradient tensor sequences to estimate µFA and other MDM parameters in individuals with recent concussion. µFA has been found to be more sensitive to WM abnormalities and less sensitive to fiber directionality than FA (Andersen et al., 2020; Ikenouchi et al., 2020; Lasič et al., 2014). MDM imaging also allows for the estimation of additional parameter distributions that capture both the mean and variance of diffusion tensor “shape” and “size” information at the subvoxel level, as opposed to simple voxel averaging (Topgaard, 2019). Broadly, MDM “shape” parameters (µFA and D^2^_anison_) correspond with the degree of directional (anisotropic) diffusion along WM tracts, while MDM “size” parameters correspond with the degree (D_iso_, also known as mean diffusivity) and normalized variability (V_ison_) of free, unrestricted (isotropic) diffusion in a voxel.

Our investigation had three aims: 1) To characterize whether and how MDM shape and size parameters are associated with acute concussion diagnosis, and, utilizing a logistic Bayesian framework, assess whether WM abnormalities as detected by these parameters manifest within a few specific regions vs. widely across the brain; 2) To compare whether MDM parameters are more sensitive to acute concussion diagnosis than FA (i.e., display better discriminability of acute concussion vs. control groups) utilizing machine learning techniques; and 3) To determine whether FA and MDM parameters are differentially associated with concussion in areas with vs. without crossing fibers, assessing their robustness to fiber directionality variation and exploring whether multidirectional voxels reflect greater differences between control and concussion groups due to shearing-type injury. We hypothesized that MDM parameters, particularly µFA, would be more sensitive to acute concussion than FA and would maintain the same relationship with acute concussion in areas with vs. without crossing fibers.

## 2. Methods

### 2.1 Concussion Group

A total of 30 participants completed all study recruitment, pre-screening, and protocol procedures (**Table 1**). Study inclusion criteria required that participants be adults between the ages of 18 – 30 (all genders, sexes, and races/ethnicities) who were recently diagnosed with concussion and could complete the study scan within five days of injury occurrence. Participants could have any history of prior head injuries. Exclusionary criteria included penetrating injury, complicated traumatic brain injury, visible signs of injury on structural MRI scan (see *Clinical MRI Scans*), and contraindications to MRI (non-removable metal, incompatible medical devices, hearing loss, tinnitus, claustrophobia), and pregnancy. While most participants had no clinically significant cardiac, neurological, pulmonary, or psychiatric diseases other than mild-moderate mood and attention deficit disorders, a few participants had other co-occurring conditions. Participants were referred to the study team by primary care practitioners at the University of California, Santa Barbara student health center. To assess eligibility, participants were asked prescreening questions based on inclusion/exclusion criteria over the phone. All participants provided written informed consent for study procedures approved by the Institutional Review Board at University of California, Santa Barbara. Participants were paid $20 for the full MRI session.

**Table 1.** Demographic Information.

|  | Concussion | Control |
| --- | --- | --- |
| N | 30 | 32 |
| Age | 21.1 (18-29)* | 22.8 (19-29)* |
| Sex | 13M/17F | 18M/14F |
| Handedness | 26R/2L<sup>+</sup> | 28R/2L/1M<sup>+</sup> |
| Previous Concussion | 11Y/19N | 6Y/26N |
| RPQ-3 | 5.8 (0-11)<sup>†</sup> | N/A |
| RPQ-13 | 22.6 (0-46)<sup>†</sup> | N/A |
| ACE Symptom Score | 12.3 (0-21)<sup>†</sup> | N/A |

Values are reported as mean (min value – max value)
\* Indicates significant difference at $p < 0.05$
<sup>+</sup>Three participants (one concussion, two control) did not complete handedness inventory
<sup>†</sup>Indicates group score is significantly greater than 0 ( $p < 0.001$ ). One concussion participant did not complete these measures.
Abbreviations: M = male, F = female, R = right, L = left, M = mixed, Y = yes, N = not reported, RPQ = Rivermead post-concussion questionnaire, ACE = Acute concussion evaluation

### 2.2 Healthy Control Group

A total of 37 young, healthy participants completed all study recruitment, pre-screening, and protocol procedures. Data from 3 participants were removed due to poor image quality, and 2 were excluded due to usage of prescribed anticonvulsant medication, leading to a total of 32 participants (**Table 1**). Study inclusion criteria required that participants be adults between the ages of 18 – 30 (all genders, sexes, and races/ethnicities). Exclusionary criteria included previous concussion diagnosis within a year prior to participation, contraindications to MRI (non-removable metal, incompatible medical devices, hearing loss/tinnitus, claustrophobia), pregnancy, or clinically significant cardiac, neurological, pulmonary, or psychiatric diseases other than mild-moderate mood (anxiety or depression) and attention deficit disorders. Prescription medications for mood and attention-deficit disorders, allergies, and birth control were allowed. Participants were recruited via word of mouth within the University of California, Santa Barbara community. To assess eligibility, participants were asked prescreening questions based on inclusion/exclusion criteria. All participants provided written informed consent for study procedures approved by the Institutional Review Board at University of California, Santa Barbara. Participants were paid $20 for the full MRI session.

### 2.3 Study Protocol

On the day of scanning, control participants completed the Edinburgh Handedness Inventory (Oldfield, 2013) and a state questionnaire assessing demographics, medication use, and health history. Concussion participants completed the same questionnaires as controls and additionally completed the Rivermead Post Concussion Inventory (King et al., 1995) and the Acute Concussion Evaluation (Gioia et al., 2008). Participants underwent magnetic-resonance imaging (MRI) in a Siemens 3T Prisma scanner with a 64-channel phased-array head/neck coil. First, high-resolution T1-weighted magnetization prepared rapid gradient echo (MPRAGE) anatomical scans were acquired (TR = 2500 ms, TE = 2.22 ms, FOV = 241 mm, T1 = 851 ms, flip angle = 7°, with 0.9 mm^3^ voxel size). Following the anatomical scan, a series of 4 spherical b-tensor (b = 0, 100 – 500, 1000, 1500 s/mm^2^; 3 diffusion directions) and 4 linear b-tensor (b = 0-500, 1000, 1500, 2000 s/mm^2^; 6, 10, 16, 30 diffusion directions) q-space trajectory encoding (QTE) diffusion sequences (Martin et al., 2020) were collected (TR = 6308 ms, TE = 80 ms, diffusion gradient amplitude = 80.0 mT/m, FOV = 230 mm, flip angle = 90°, 2.0 x 2.0 mm^2^ in-plane resolution, 4.0 mm slice thickness, iPAT factor = 2).

### 2.4 Clinical MRI Scans

All participants additionally underwent a battery of routine clinical brain MRI scans, identical to those used by local diagnostic radiology centers. These scans include a T_2_-weighted turbo spin echo (TSE) sequence (TR = 6000 ms, TE = 100 ms, FOV = 220 mm, flip angle = 150°, slice thickness = 4.0 mm, 0.4 x 0.4 mm^2^ in plane resolution), a T_2_-weighted FLAIR sequence (TR = 9000 ms, TE = 83.0 ms, FOV = 220 mm, flip angle = 150°, slice thickness = 4.0 mm, 0.7 x 0.7 mm^2^ in plane resolution), a T_2_*-susceptibility weighted sequence (TR = 738.0 ms, TE = 19.90 ms, FOV = 220 mm, flip angle = 20°, slice thickness = 4.0 mm, 0.7 x 0.7 mm^2^ in plane voxel size), and a diffusion imaging sequence (TR = 4100 ms, TE = 81 ms, diffusion gradient amplitude = 80.0 mT/m, FOV = 230 mm, flip angle = 90°, 2.0 x 2.0 mm^2^ in-plane resolution, 4.0 mm slice thickness, iPAT factor = 2, 3 diffusion directions). Clinical MRI scans were read blinded to group membership by a licensed neurologist with expertise in neuroimaging. No participant had evidence of mass effect, micro- or macroscopic hemorrhage, diffusion abnormalities suggestive of ischemia, or other significant findings. Nine of the control participants and ten of the concussion patients had sparse (fewer than 4), small (less than 3mm) enlarged perivascular spaces, most commonly around penetrating vessels in the superior frontal white matter and of uncertain clinical significance.

### 2.5 MRI Preprocessing

MRI preprocessing was conducted with Advanced Normalization Tools (ANTs; Avants et al., 2011) and MATLAB (The MathWorks Inc, 2018). First, T_1_-weighted anatomical data were skull-stripped with antsBrainExtraction.sh. In order to create individualized participant white matter (WM) and grey matter (GM) tissue masks, we first segmented the skull-stripped anatomical data using the ANTsPy (https://github.com/ANTsX/ANTsPy) kmeans_segmentation function, which outputted probability maps of participant WM, GM, and cerebrospinal fluid (CSF). We then binarized these probability maps (all values > 10% = 1) to obtain WM, GM, and CSF tissue masks.

Second, for each participant, we calculated region-of-interest (ROI)-specific mean values of FA and four MDM parameters, which describe facets of diffusion tensor size and shape (for parameter descriptions, see *MDM Parameters*). To do this, we first used an open source, MATLAB pipeline for MDM MRI (https://github.com/markus-nilsson/md-dmri) to complete motion and eddy current correction of diffusion data, as well as estimation of voxel-wise brain maps of size-shape-orientation diffusion tensor distributions (DTDs) using the “dtd” method (Nilsson et al., 2018; Topgaard, 2019).

Then, using ANTs, we spatially normalized participant DTD diffusion data, HCP1065 Population-Averaged Tractography Atlas probabilistic WM region masks (thresholded at ≥ 50%; Yeh, 2022), and deterministic trackable direction masks denoting fiber directionality (Volz et al., 2018) to their participant-specific anatomical space, where all further calculations were conducted. Next, we utilized the participant-specific WM tissue masks to extract white matter voxels from DTD data. Each white matter voxel was assigned 2 independent labels: one indicating its respective HCP1065 region and one indicating crossing fiber directionality. Fiber directionality categories included 0-Cross voxels (i.e., voxels with unidirectional fiber bundles, consisting of parallel fibers) and 1-Cross voxels (i.e., voxels with two-directional diffusion, indicating a single crossing of two bundles). Voxels without a crossing fiber ID (did not align to masks) and with more than 1 crossing (small sections of WM tissue have 2-Cross voxels, typically along the gray/white matter boundary) were excluded from all further analyses.

Then, for each of the 64 white matter (WM) regions, we calculated median values of all diffusion parameters. Additionally, within each region, we calculated separate median values for segments categorized by crossing fiber directionality (e.g., separate median values for corpus callosum body 0-Cross voxels and 1-Cross voxels). These segmented regions, which combine region and fiber directionality information, are here on referred to as region-cross ROIs. Only region-cross ROIs with at least 10 voxels per participant were kept for future analyses (not all participants have voxels for each region-cross combination due to individual differences in morphology).

### 2.6 MDM Parameters

To capture detailed changes in WM architecture due to acute concussion, we estimated voxel maps of four multidimensional diffusion (MDM) parameters. Two of these parameters describe diffusion tensor “shape” (**Fig 1A**; micro fractional anisotropy; μFA, normalized mean squared anisotropy; D^2^_anison_), while two describe diffusion tensor “size” (**Fig 1B**; mean isotropic diffusivity; D_iso_, normalized variation in isotropic diffusivity; V_ison_). In addition, we also estimated parameter maps of conventional FA to compare to these MDM parameters.

**Figure 1.**
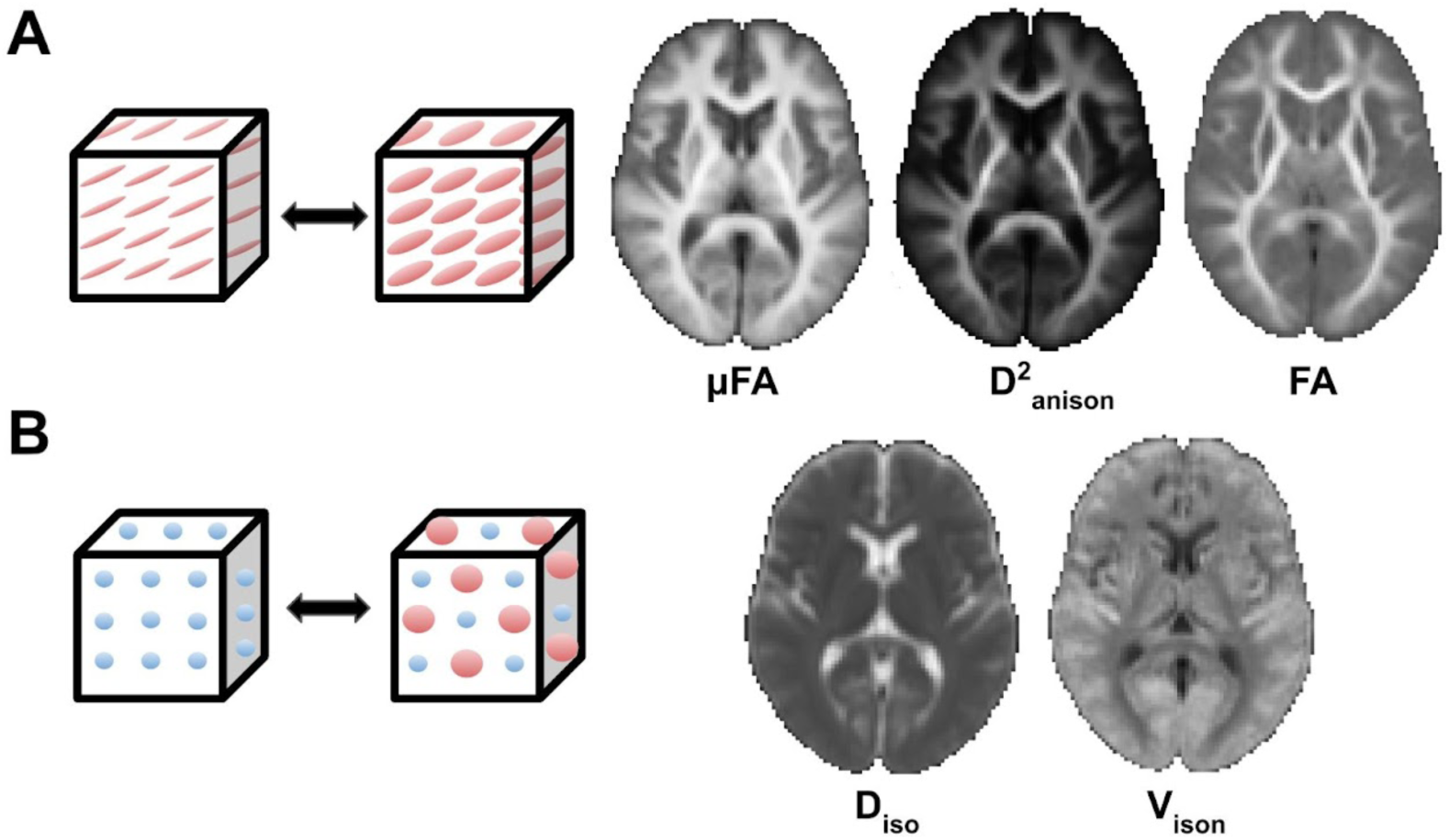
MDM and FA parameter maps averaged across all healthy control participants (N=32). **A (top row):** MDM shape parameters represent degree of anisotropic diffusion in a voxel. Higher values indicate greater anisotropic diffusion (left cube, thinner ellipses), while lower values indicate less anisotropic diffusion (right cube, thicker ellipses). While all 3 shape parameters measure similar properties, μFA (left) remains high in WM regions with crossing fibers, while FA (right) decreases. **B (bottom row):** MDM size parameters represent degree (D_iso_) and variability (V_ison_) of isotropic diffusion in a voxel. Greater values indicate greater degree (D_iso_; larger spheres) and variability (V_ison_; more varied spheres) of isotropic diffusion (right cube), while lower values indicate less degree and variability of isotropic diffusion (left cube, smaller uniform spheres). For all parameter maps, brighter areas have greater values, while darker areas have lower values.

As defined by Topgaard, 2019, we first estimated distributions of parallel (D_||_) and perpendicular (D_⊥_) component diffusivities (in 10^-8^ m^2^/s) within each voxel. Mean isotropic diffusivity (D_iso_) and normalized diffusion anisotropy (D_anison_, represented in Topgaard, 2019 as D_Δ_) were calculated as shown below (Topgaard, 2019, equation 2):

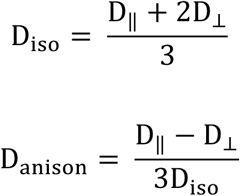

We then took the expected values (E[x]) of D_iso_ and D_anison_ to obtain E[D_iso_], which is equivalent to mean diffusivity (MD), and E[D^2^_anison_], which contains similar information as other measures of tensor anisotropy (Topgaard, 2019, equations 9 and 10), as shown below:

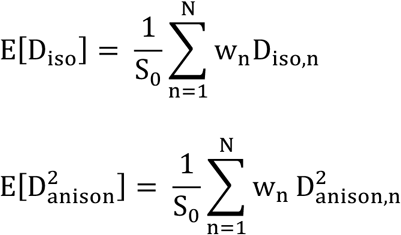

To obtain V_ison_, we calculated the variance of isotropic diffusivity (Var[D_iso_]; Topgaard, 2019, equation 11) normalized by D^2^_iso_, as shown below:

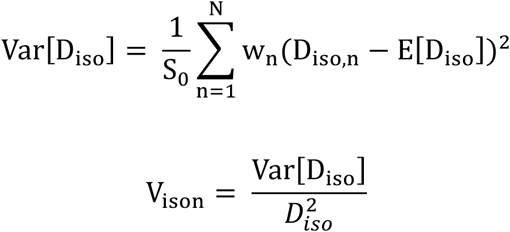

μFA was calculated according to equation 14 in Lasič et al., 2014:

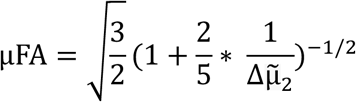

Where Δµ<_-_ is “the measurable difference in curvature between powder-averaged and isotropic signal-vs.-b data” (Lasič et al., 2014, equation 13):

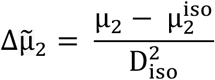

As discussed in Lasič et al., 2014, equation 22, FA can be derived from μFA and the orientation dispersion parameter (OP), also known as “microscopic orientation coherence”, or the ratio of micro- to macroscopic anisotropy in a voxel. Participant μFA and OP data were used to calculate their respective FA maps:

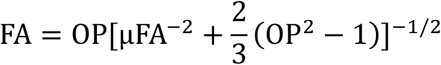

OP was calculated according to equation 33 in Westin et al., 2016 (here, OP is represented as C_c_):

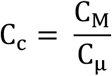

Where C_M_ is the estimated macroscopic anisotropy, while Cμ is the microscopic anisotropy. The orientation parameter ranges from 0 (diffusion in all directions equally, whether it be isotropic or anisotropic) to 1 (parallel diffusion in one direction). Both μFA and FA range from 0 (completely isotropic diffusion) to 1 (completely anisotropic diffusion); however, in voxels with crossing fibers (lower OP), μFA retains higher values (reflecting multidirectional anisotropic diffusion), while FA values decrease (Lasič et al., 2014).

### 2.7 Statistical Analyses

All analyses were conducted using Python (version 3.11.5). We first aimed to assess whether concussion and control groups differed in age, and whether concussion group symptom scores (RPQ-3, RPQ-13, ACE) were significantly different from zero. To do this, one two-sided t-test (age) and three one-sided t-tests (symptom scores) using the pingouin (0.5.3) package were conducted.

Next, we tested whether FA and any of the 4 MDM parameters are associated with concussion diagnosis with Bayesian univariate Gaussian mixture logistic regression models. Ten total logistic regression models were run to predict the log-odds of a participant’s group membership (μ) based on their MDM diffusion parameter values while controlling for age and sex. Five models (corresponding to 5 diffusion measures [D_iso_, V_ison_, D_anison_, μFA, FA]) are here on referred to as the **Region Model,** where, for each WM whole region, a single beta coefficient (β_r_) corresponding to its median diffusion parameter value was fitted across participants:

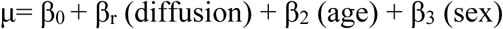

The other five models are here on referred to as the **Crossing Fiber Model,** where, for each WM region-cross ROI (e.g., all 0-Cross voxels within corpus callosum tapetum), a single beta coefficient (β_r,c_) corresponding to its median diffusion parameter value was fitted across participants in each group:

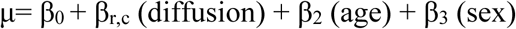

Median diffusion parameter and participant age values were z-score normalized prior to entry into the model. In order to estimate likelihood of group membership, μ was transformed with a sigmoid function to obtain probability for a Bernoulli likelihood function.

We sampled β posterior distributions using No U-Turn sampling (NUTS) Hamiltonian Monte Carlo, implemented with the PyMC package (Salvatier et al., 2016). Posteriors were sampled in four parallel chains of 5000 samples (20000 total) with an additional initial 5000 samples per chain (tuning samples were then discarded). We required that no chain contain any divergences. We then calculated highest density intervals of the diffusion beta posterior distributions (highest density intervals [HDIs]; the Bayesian equivalent of a confidence interval) using 94% density in the arviz package (Kumar et al., 2019). In all tables, β_r_ and β_r,c_ values are reported as the median of its estimated posterior distribution. Odds ratios are reported as the median of the exponentiated posterior distribution. Credible distributions are reported as those where the 94% HDI does not contain zero.

Finally, to directly compare group discriminability of MDM parameters vs. that of FA, we employed a stratified k-fold cross-validated logistic regression model with scikit-learn in Python, from here on referred to as the **Region Classifier Model**. As with previously described models, each logistic regression model aimed to predict concussion vs. control group membership in a specific WM region (whole region, not split according to fiber directionality) based on that region’s diffusion parameter values while controlling for participant age and sex. Median diffusion parameter and participant age values were z-score normalized prior to entry into the model. Briefly, for a given WM region and diffusion parameter combination, participant data were split into k=4 folds; at each iteration, a logistic regression model (LBFGS solver with a maximum of 300 iterations) was fit to the training set (3 folds) and predicted probabilities of concussion group membership on the left-out test set (1 fold). Data were stratified such that each fold had a roughly equal amount of concussion and control participants. Predicted probabilities across all folds were then aggregated, and model performance for that WM region (closeness of predicted vs. true values) was evaluated by calculating the area under a receiver operator characteristic curve (AUC). This process was performed such that we had 320 AUC values (64 regions x 5 diffusion parameters). Finally, to compare model performance (AUC) between each of 4 MDM parameters and FA, we performed 4 within-region t-tests (scipy ttest_rel); each t-test compared the regional distribution of AUC values for a given MDM parameter vs. the equivalent regional distribution of AUC values for FA. Significance was evaluated with a Bonferroni adjusted p-value of p<0.0125 (p=0.05/4 comparisons).

## 3. Results

### 3.1 Demographics

We first assessed whether there existed demographic differences between concussion and control groups, and whether the concussion group verifiably showed presence of symptoms as assessed via symptom questionnaires. T-test analyses revealed a significant difference in age between the groups (p<0.05) and that the concussion group’s symptom scores across three questionnaires were significantly different than zero (p<0.001; **Table 1**). Because the groups showed a significant difference in age, we controlled for age and sex in all models. First, we explored whether MDM parameters and FA across WM regions are associated with presence of acute concussion (“Region Model”). Then, we assessed MDM parameter group discriminability vs. that of FA (“Region Classifier Model”) by computing classifier performance at each region (AUC values) and comparing MDM-to-FA AUC (see **Methods**). Finally, we explored whether MDM parameter and FA associations with acute concussion vary across subregions with vs. without crossing fibers (“Crossing Fiber Model”).

### 3.2 Region Model: MDM parameters are associated with acute concussion

We found that all MDM parameters and FA were credibly associated with acute concussion in at least one region, regardless of whether they measured diffusion tensor “shape” (anisotropic diffusion; μFA, D^2^_anison_), or degree (D_iso_) or variability (V_ison_) of diffusion tensor “size” (isotropic diffusion).

Acute concussion diagnosis was associated with lower values of FA as well as the MDM shape parameters μFA and D^2^_anison_ (i.e., less regional anisotropic diffusion along WM tracts, **Table 2**). FA was only credibly associated with concussion in one region, while μFA was credibly associated in five regions. D^2^_anison_ was credibly associated with concussion in more regions than all other shape parameters (nine regions; **Fig 2**). All credible regions for shape parameters had negative β_r_ values, indicating that concussion was consistently associated with less anisotropic diffusion regardless of the region examined.

**Table 2.** Region Model: Credible logistic regression β posteriors.

| Region Name | $\beta_r$ | HDI | Odds Ratio |
| --- | --- | --- | --- |
| **FA** |  |  |  |
| Cingulum Parahippocampal (R) | -1.818 | [-3.726, -0.018] | 0.162 |
| **μFA** |  |  |  |
| Corticobulbar Tract (L) | -5.308 | [-9.861, -1.095] | 0.005 |
| Corticobulbar Tract (R) | -4.834 | [-9.18, -0.788] | 0.008 |
| Corticopontine Tract Frontal (R) | -5.283 | [-10.499, -0.639] | 0.005 |
| Corticostriatal Tract Superior (R) | -4.726 | [-9.105, -0.437] | 0.009 |
| Reticulospinal Tract (L) | -4.648 | [-9.5, -0.043] | 0.010 |
| **D<sup>2</sup><sub>anison</sub>** |  |  |  |
| Cingulum Rarolfactory (L) | -1.557 | [-3.162, -0.09] | 0.211 |
| Corticobulbar Tract (L) | -1.747 | [-3.219, -0.317] | 0.174 |
| Corticobulbar Tract (R) | -1.706 | [-3.207, -0.29] | 0.182 |
| Corticopontine Tract Frontal (L) | -1.688 | [-3.376, -0.126] | 0.185 |
| Corticopontine Tract Frontal (R) | -2.053 | [-3.889, -0.352] | 0.128 |
| Corticospinal Tract (L) | -1.548 | [-3.172, -0.049] | 0.213 |
| Corticospinal Tract (R) | -1.489 | [-3.08, -0.069] | 0.226 |
| Corticostriatal Tract Superior (R) | -2.269 | [-4.248, -0.472] | 0.103 |
| Reticulospinal Tract (L) | -2.235 | [-4.489, -0.14] | 0.107 |
| **D<sub>iso</sub>** |  |  |  |
| Corpus Callosum Tapetum | 1.606 | [0.124, 3.354] | 4.982 |
| Corticostriatal Tract Anterior (R) | 2.866 | [0.279, 5.63] | 17.563 |
| **V<sub>ison</sub>** |  |  |  |
| Cingulum Frontal Parahippocampal (L) | 1.490 | [0.131, 2.968] | 4.437 |
| Cingulum Frontal Parietal (L) | 1.479 | [0.019, 2.991] | 4.390 |
| Corpus Callosum Forceps Major | 2.613 | [0.533, 4.744] | 13.637 |
| Corticopontine Tract Frontal (L) | 2.017 | [0.375, 3.937] | 7.513 |
| Corticopontine Tract Occipital (L) | 1.827 | [0.524, 3.274] | 6.218 |
| Corticospinal Tract (L) | 2.244 | [0.27, 4.543] | 9.431 |
| Corticostriatal Tract Posterior (L) | 2.181 | [0.492, 4.097] | 8.851 |
| Fornix (L) | 1.012 | [0.137, 1.928] | 2.750 |
| Inferior Fronto Occipital Fasciculus (L) | 2.150 | [0.263, 4.106] | 8.586 |
| Inferior Longitudinal Fasciculus (L) | 3.563 | [1.31, 6.034] | 35.253 |
| Inferior Longitudinal Fasciculus (R) | 1.775 | [0.004, 3.658] | 5.899 |
| Optic Radiation (L) | 1.671 | [0.362, 3.074] | 5.316 |
| Optic Radiation (R) | 1.422 | [0.188, 2.756] | 4.145 |
| Reticulospinal Tract (L) | 1.147 | [0.076, 2.395] | 3.150 |
| Superior Longitudinal Fasciculus1 (R) | 1.702 | [0.169, 3.354] | 5.487 |
| Thalamic Radiation Anterior (L) | 1.157 | [0.075, 2.35] | 3.180 |
| Thalamic Radiation Posterior (L) | 1.808 | [0.195, 3.66] | 6.100 |
| Thalamic Radiation Superior (L) | 1.974 | [0.116, 4.067] | 7.200 |
| Uncinate Fasciculus (R) | 0.937 | [0.085, 1.843] | 2.553 |
| Vertical Occipital Fasciculus (R) | 1.434 | [0.14, 2.848] | 4.197 |

**Figure 2.**
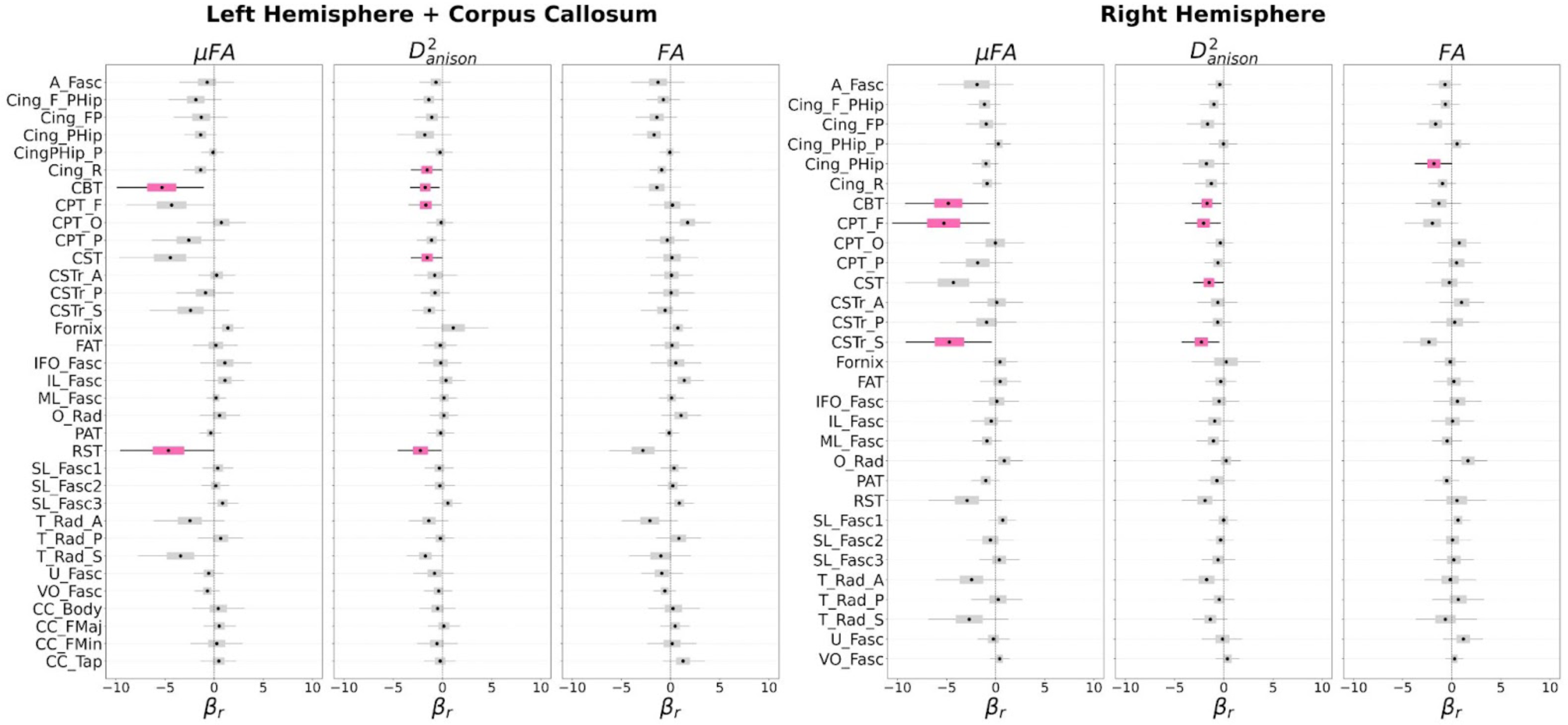
Region Model posteriors for MDM tensor shape parameters. Credible β_r_ posteriors (94% HDI does not contain zero) are bolded and shaded pink. All credible regions have negative β_r_ values, which indicate that acute concussion is more likely (higher log odds) with lower values of that parameter in that region. **Abbreviations: A_Fasc**: Arcuate Fasciculus, **Cing_F_PHip**: Cingulum Frontal Parahippocampal, **Cing_FP**: Cingulum Frontal Parietal, **Cing_PHip**: Cingulum Parahippocampal, **CingPHip_P**: Cingulum Parahippocampal Parietal, **Cing_R**: Cingulum Rarolfactory, **CBT**: Corticobulbar Tract, **CPT_F**: Corticopontine Tract Frontal, **CPT_O**: Corticopontine Tract Occipital, **CPT_P**: Corticopontine Tract Parietal, **CST**: Corticospinal Tract, **CSTr_A**: Corticostriatal Tract Anterior, **CSTr_P**: Corticostriatal Tract Posterior, **CSTr_S**: Corticostriatal Tract Superior, **Fornix**: Fornix, **FAT**: Frontal Aslant Tract, **IFO_Fasc**: Inferior Fronto Occipital Fasciculus, **IL_Fasc**: Inferior Longitudinal Fasciculus, **ML_Fasc**: Middle Longitudinal Fasciculus, **O_Rad**: Optic Radiation, **PAT**: Parietal Aslant Tract, **RST**: Reticulospinal Tract, **SL_Fasc1**: Superior Longitudinal Fasciculus1, **SL_Fasc2**: Superior Longitudinal Fasciculus2, **SL_Fasc3**: Superior Longitudinal Fasciculus3, **T_Rad_A**: Thalamic Radiation Anterior, **T_Rad_P**: Thalamic Radiation Posterior, **T_Rad_S**: Thalamic Radiation Superior, **U_Fasc**: Uncinate Fasciculus, **VO_Fasc**: Vertical Occipital Fasciculus, **CC_Body**: Corpus Callosum Body, **CC_FMaj**: Corpus Callosum Forceps Major, **CC_FMin**: Corpus Callosum Forceps Minor, **CC_Tap**: Corpus Callosum Tapetum

In contrast, acute concussion diagnosis was associated with greater values of the size parameters D_iso_ and V_ison_ (i.e., more regional mean and variation in isotropic diffusion or freely diffusing water, **Table 2**). More specifically, D_iso_ was credibly associated with concussion in only two regions, while V_ison_ was credibly associated in 20 (**Fig 3**). Notably, V_ison_ was credibly associated in many regions not observed for the shape parameters, such as thalamic radiation and longitudinal fasciculus tracts. All credible regions for size parameters had positive β_r_ values, indicating that concussion was consistently associated with greater and more variable isotropic diffusion regardless of the region examined.

**Figure 3.**
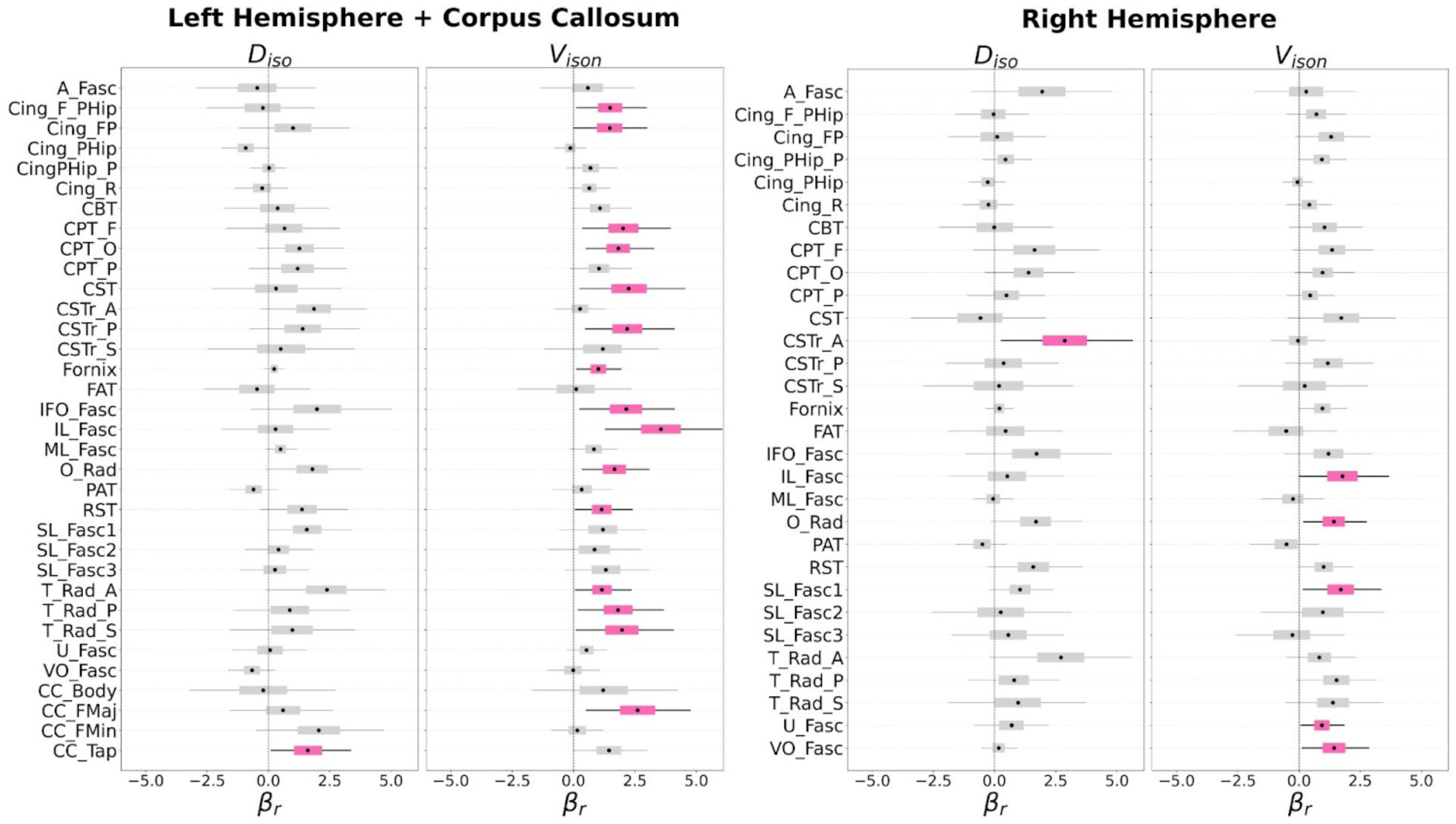
Region Model posteriors for MDM tensor size parameters. Credible β_r_ posteriors (94% HDI does not contain zero) are bolded and shaded pink. All credible regions have positive β_r_ values, which indicate that acute concussion is more likely (higher log odds) with greater values of that parameter in that region.

Overall, both shape (negative association) and size (positive association) MDM parameters showed consistent relationships with acute concussion (directionality did not vary based on the region examined). Parameters were credible among a wide variety of regions; yet, a subset of regions were consistently credible across parameters, including corticopontine, corticostriatal, corticospinal, and reticulospinal tracts.

### 3.3 Region Classifier Model: MDM shape parameters more sensitive to acute concussion than FA

After characterizing MDM parameter associations with concussion, we directly compared whether these parameters are more sensitive to acute concussion (better group discrimination) than FA across WM regions (**Table 3**). Both MDM shape parameters (μFA and D^2^_anison_) had significantly greater regional AUC values than FA (p<0.001; **Fig 4**), indicating better discriminability. On the other hand, both MDM size parameters D_iso_ and V_ison_ also had significantly greater AUC values than FA (p<0.05; **Fig 4**), but did not pass multiple comparison correction. Overall, only MDM shape parameters were found to be significantly more sensitive to acute concussion than FA.

**Table 3.** Region Classifier Model: MDM vs. FA Classifier Performance.

| Diffusion Parameter | AUC | MDM vs. FA p-value |
| --- | --- | --- |
| FA | 0.65 (0.56-0.71) | N/A |
| μFA | 0.66 (0.61-0.72) | μFA > FA; $p < 0.001$ |
| $D^2_{\text{anison}}$ | 0.67 (0.61-0.72) | $D^2_{\text{anison}} > \text{FA}$; $p < 0.001$ |
| $D_{\text{iso}}$ | 0.66 (0.59-0.71) | $D_{\text{iso}} > \text{FA}$; $p = 0.026$ |
| $V_{\text{ison}}$ | 0.66 (0.59-0.77) | $V_{\text{ison}} > \text{FA}$; $p = 0.028$ |

AUC values are reported as mean (min value - max value)

**Figure 4.**
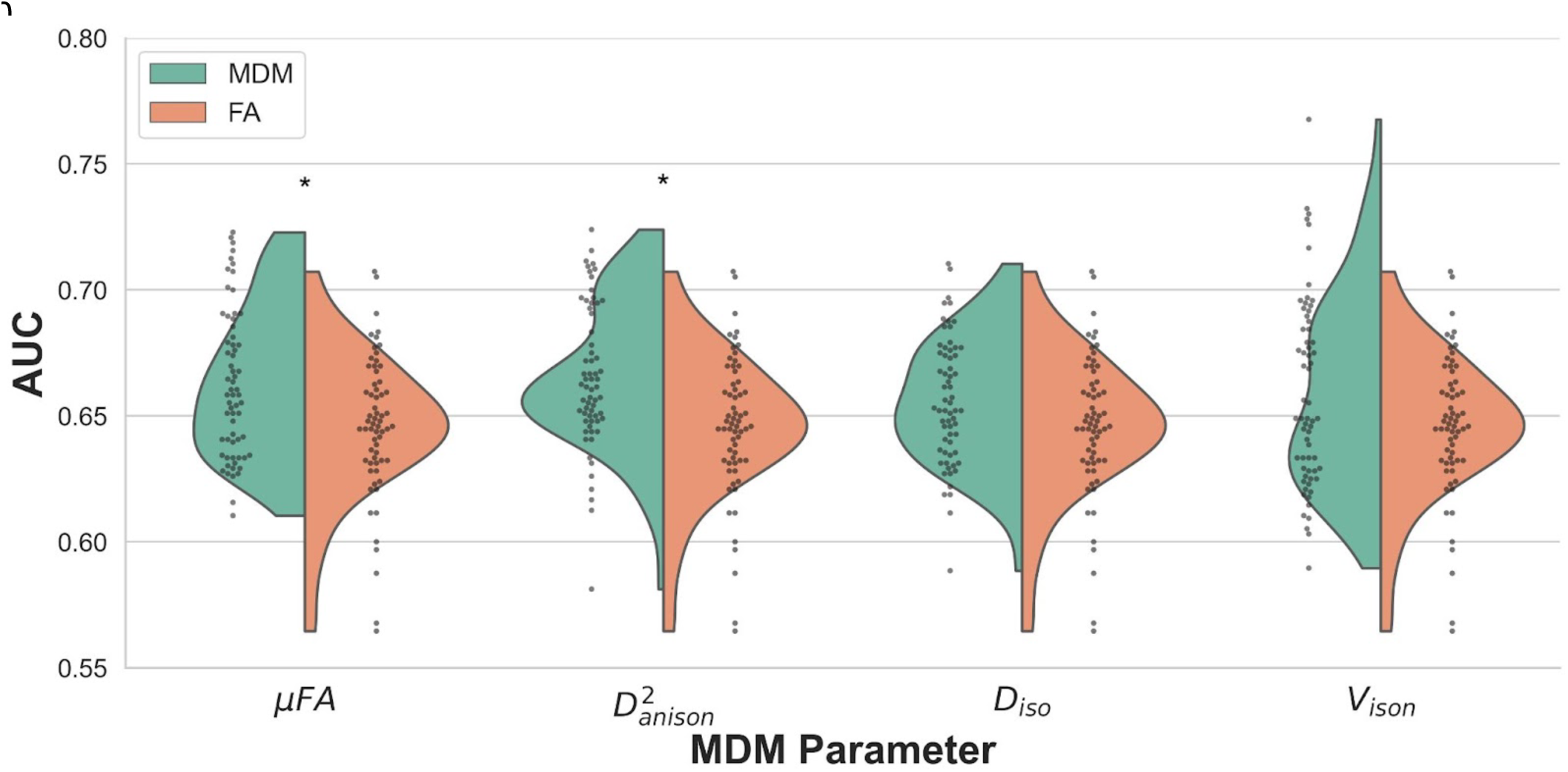
MDM Parameter vs. FA Classifier Performance. For all WM regions (represented as points), regional MDM AUC (classifier performance) was evaluated against FA AUC. * indicates significant difference in regional AUC distributions between the MDM parameter (green distribution) and FA (orange distribution). MDM shape parameters were found to have better concussion vs. control group discriminability (significantly greater AUC values) than FA (p<0.001).

### 3.4 Crossing Fiber Model: MDM parameters are associated with acute concussion in subregions with and without crossing fibers

Finally, we assessed whether MDM parameter associations with concussion differ according to fiber directionality. The Crossing Fiber Model was fit with the same Bayesian logistic regression framework as the Region Model, but with WM region-cross ROIs as opposed to whole regions. WM region-cross ROIs are either 0-Cross subregions that contain unidirectional voxels (parallel fiber bundles) or 1-Cross subregions that contain multidirectional voxels (two crossing fiber bundles). MDM parameters and FA were credibly associated with acute concussion in at least one region-cross ROI (**Table S1**).

As observed in the Region Model, lower values of MDM shape parameters (μFA and D^2^_anison_) were credibly associated with acute concussion regardless of the region. The Crossing Fiber model additionally revealed that this trend remained stable regardless of fiber directionality (**Fig S1**). Notably, FA was only credibly associated with concussion in 2 region-cross ROIs and relationships did vary based on fiber directionality (0-Cross left parahippocampal cingulum: negative; 1-Cross left inferior fronto-occipital fasciculus: positive). MDM size parameters also mimicked Region Model trends (**Fig S2**), where greater values of D_iso_ and V_ison_ were associated with acute concussion. The Crossing Fiber Model additionally revealed that these relationships were largely invariant to fiber directionality. The only exception lies in the 0-Cross ROI (parallel fibers) of the left parahippocampal cingulum, where concussion was associated with lower values of D_iso_.

Overall, defining subregions by crossing fiber directionality did not meaningfully alter the relationships between MDM parameters and acute concussion observed in the Region Model. Credible association with acute concussion was observed in both 0-Cross and 1-Cross tracts across parameters, suggesting that concussion-induced WM abnormalities may be invariant to fiber directionality.

## 4. Discussion

In the current study, we tested whether MDM diffusion parameters were credibly associated with acute concussion diagnosis. Overall, we found that the log-odds of acute concussion diagnosis increased with reduced values of MDM shape parameters (μFA, D^2^_anison_) and elevated values of MDM size parameters (D_iso_, V_ison_; **Fig 5**), with patterns persisting across several WM regions. Notably, we also identified that MDM shape parameters were significantly better at discriminating between concussion and control groups than FA. Finally, we found that for μFA, D^2^_anison_, and V_ison_, relationships with acute concussion were invariant of fiber directionality (i.e., patterns remained largely unchanged in subregions with and without crossing fibers), while FA and D_iso_ differed.

**Figure 5.**
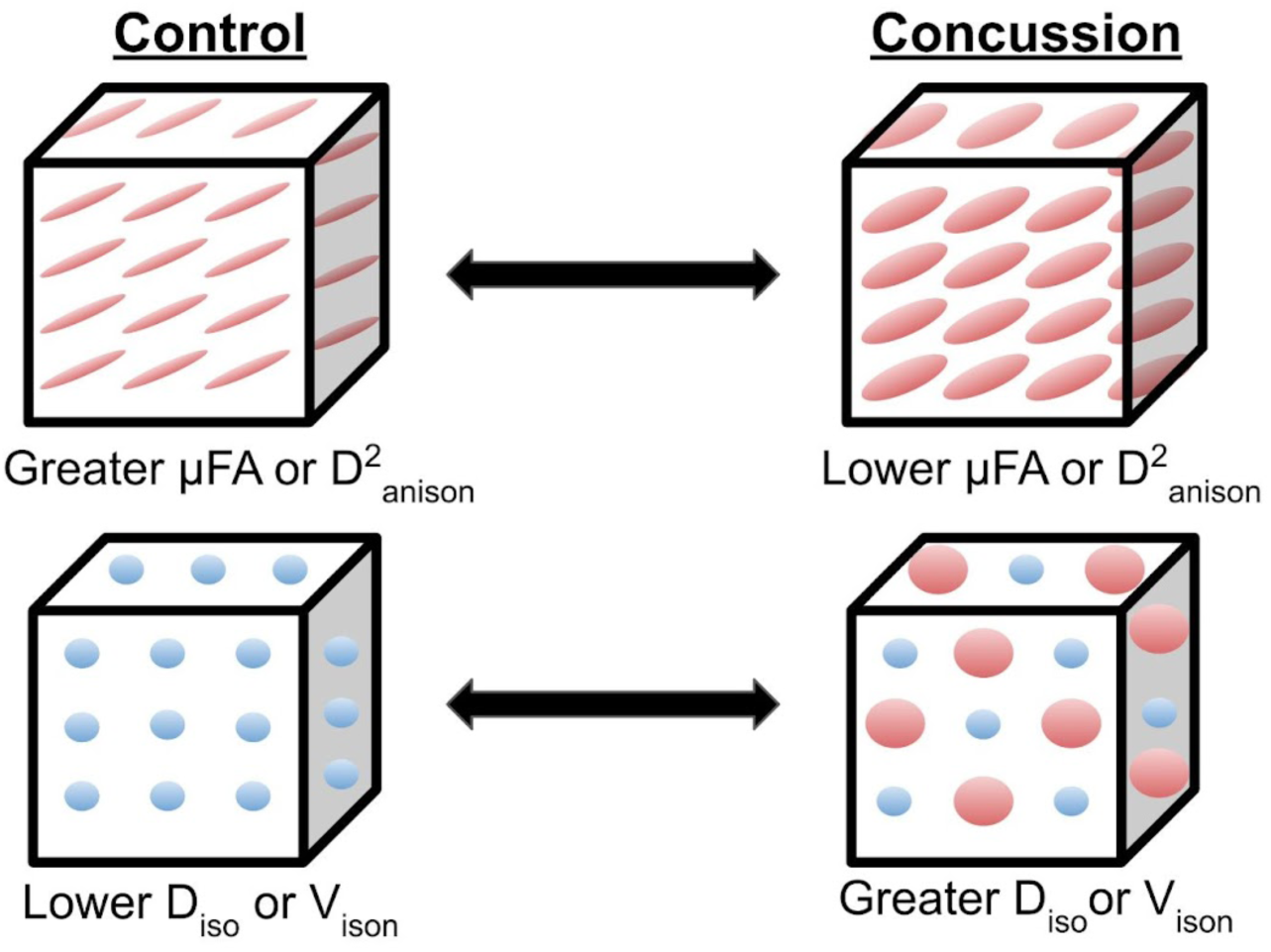
Summary of MDM parameter-concussion associations. Our MDM imaging results suggest the following model to describe WM injury after concussion. Top row: Water typically moves along the direction of the WM fibers (left). After a head impact (concussion), water is less restricted in its usual direction of movement (right), as reflected by lower values of MDM shape parameters. Bottom row: Free movement of water is typically constrained in areas between WM fibers (left). After concussion, water moves more freely in these tiny areas (right, big red spheres), and there is more variability in the amount of this movement (big vs. small spheres), as reflected by greater values of MDM size parameters. Our results do not show exactly what causes these changes, but capturing these changes may be useful for detecting subacute concussion. Figure design is borrowed from Topgaard, 2019.

Our results suggest that MDM shape and size parameters can serve as biomarkers of acute concussion and that these biomarkers signal abnormalities in widespread areas of the brain. Concussion was associated with lower values of μFA and D^2^_anison_ (lower diffusion anisotropy); not surprisingly, these indicators were credible in many similar regions, as both parameters capture similar information about structural anisotropy (Topgaard, 2017). Future work is needed to uncover whether our observed decreases in μFA and D^2^_anison_ reflect known concussion-induced microstructural changes, such as axonal demyelination (Mahoney et al., 2022; Shi et al., 2015). On the other hand, concussion was associated with greater values of D_iso_, the MDM equivalent to mean diffusivity (MD), conflicting with previous DTI work that typically found decreased MD in acute concussion (Henry et al., 2011; Hulkower et al., 2013; Lancaster et al., 2016). Our other size parameter, V_ison_, is the variance of D_iso_, or the variability of the degree of freely diffusing water within a voxel (MD). While MD (D_iso_) has been often examined in concussion, MD variance has been less considered due to interpretive challenges and unavailability of the measure through conventional DTI. The current study revealed that acute concussion diagnosis was also associated with greater values of V_ison_. Similarly, preliminary work with MDM parameters found increased D_iso_ (MD) and V_ison_ (MD variance) in the WM of chronic schizophrenia patients (Westin et al., 2016) potentially reflecting neuroinflammatory increases in extracellular free water (Pasternak et al., 2012). Evidence also exists that V_ison_ positively correlates with cell density heterogeneity in brain tumors (validated with quantitative microscopy; Szczepankiewicz et al., 2016). Although the observed increases in V_ison_ here might indicate concussion-induced changes in cell density; future work is needed to clarify whether any post-injury neuroinflammatory processes, such as a cellular infiltrate, are reflected by MD variance measures.

We identified abnormal FA in only two regions (inferior fronto-occipital fasciculus and parahippocampal cingulum); these areas were also identified in a recent meta-analysis of acute concussion (Hellewell et al., 2020). However, we did not observe FA abnormalities in commonly identified sites across DTI studies, such as the corpus callosum and the internal capsule (Hulkower et al., 2013; Niogi et al., 2008; Wilde et al., 2008). Conversely, our MDM shape and size parameters were credible in many regions across the brain, including callosal tracts and projection tracts that pass through the internal capsule. V_ison_ was credible within association fibers not observed for other parameters, such as thalamic radiation and longitudinal fasciculus tracts. Thus, assessing V_ison_ may be a promising approach for characterizing widespread abnormalities across many types of WM tracts. Because our concussion group population varied widely in injury characteristics (see **Supplementary Table S3**), our group results may reflect widespread neuroinflammatory processes invariable to regional location of impact. Longitudinal, within-subject approaches are needed to identify whether concussion-induced effects manifest more severely in specific regions based on individual injury characteristics.

After demonstrating that MDM parameters can potentially serve as biomarkers of acute concussion, we found that shape parameters μFA and D^2^_anison_ were better at discriminating between acute concussion vs. control groups than FA. μFA has the intuitive advantage of being similar to the widely used FA, with the added advantage of controlling for subvoxel variation in fiber orientation (i.e., crossing, kissing and fanning fibers). Prior studies have demonstrated the value of μFA for assessing anisotropy in the contexts of multiple sclerosis (Andersen et al., 2020), Parkinson’s disease (Ikenouchi et al., 2020), temporal lobe epilepsy (Arezza et al., 2024), discrimination of brain tumor tissue (Szczepankiewicz et al., 2016), and as a biomarker of brain structural changes across the menstrual cycle (Rizor et al., 2024). While the size parameters D_iso_ (MD) and V_ison_ (MD variance) showed greater discriminability than FA, the differences observed were not statistically significant. Debate exists over whether MD or FA are preferable biomarkers of concussion; recent work found that MD outperformed FA as a classifier of concussion diagnosis (Ly et al., 2022). Our results suggest that enhanced measures of anisotropy (i.e., those less sensitive to fiber orientation) may be more reliable for classifying brain injury than FA or conventional measures of isotropy (MD).

Our final aim was to determine whether WM abnormalities as detected by MDM parameters manifest differently across tissue of varying fiber directionality. Previous DTI studies have generated conflicting FA results that may reflect FA’s sensitivity to fiber orientation (Hellewell et al., 2020). Similarly, we observed that FA’s relationship with presence of concussion differed based on fiber directionality (i.e., negative association in a 0-Cross region and positive association in a 1-Cross region). This discrepancy highlights the advantage of utilizing μFA, which demonstrated consistent negative relationships with concussion regardless of region or fiber directionality. We additionally observed that concussion-induced WM abnormalities were not exclusively present in WM regions with multidirectional fibers. While shearing-related damage (more likely in multidirectional tissue) has been identified in more severe traumatic brain injury (Johnson et al., 2013; Strich, 1961) with regard to concussion, we detected widespread diffusion changes throughout the white matter regardless of fiber directionality. This may be because our concussion group experienced injuries too mild to cause shearing-specific effects, or our parameters were not uniquely sensitive to these effects, instead detecting more widespread phenomena.

The current study has limitations that should be considered for future research approaches. More concussion group members reported previous head injuries than the control group; thus, our results could partially reflect lingering chronic concussion effects. Secondly, our study was cross-sectional, potentially obscuring individual differences in injury effects. The between-group design also allows for potential group differences unrelated to concussion to influence parameter associations with group membership. Future within-subject, longitudinal studies with MDM parameters are warranted to assess how these parameters may reflect neuroanatomical changes in WM injury recovery. Additionally, future large cohort studies are needed to determine more accurate sensitivity and specificity metrics for MDM parameters. Other advancements in diffusion imaging techniques, such as neurite orientation dispersion and density imaging (NODDI), diffusion kurtosis imaging (DKI), and fixel-based analyses have allowed for parameters with greater microstructural specificity than FA (Jensen et al., 2005; Raffelt et al., 2015; Zhang et al., 2012). It is unclear which imaging methodologies are best for capturing biomarkers of concussion-induced injury. MDM parameters have been shown to provide greater specificity than DKI, as DKI entangles size and shape parameters and does not allow for the estimation of an MD variance measure (Topgaard, 2017; Westin et al., 2016). Both NODDI and fixel-based analyses have shown promise for characterization of post-concussion WM injury, though NODDI’s axial diffusivity measure assumes uniform fiber orientation (Howard et al., 2022; Mito et al., 2022; Palacios et al., 2020). Future investigations should compare MDM parameter performance to other imaging modalities; applying parameter combinations across imaging modalities may be ideal for characterizing the complicated “neurometabolic cascade” of effects experienced post-concussion (Giza & Hovda, 2014).

## Data and Code Availability

Upon publication, all study MRI data will be freely available at https://openneuro.org/datasets/ds006445. All code for analyses will be available at https://github.com/ejrise/mdm_con.

## Author Contributions

**Elizabeth J. Rizor:** Conceptualization, Methodology, Formal Analysis, Investigation, Data Curation, Writing-Original Draft, Visualization. **Neil M. Dundon:** Methodology, Formal Analysis, Writing – Review & Editing, Visualization. **Margaret Hayes:** Investigation, Project Administration. **Kiana Sabugo:** Investigation, Project Administration. **Scott T. Grafton:** Conceptualization, Methodology, Resources, Writing – Review & Editing, Supervision, Funding Acquisition

## Funding

This research was supported by the Institute for Collaborative Biotechnologies under Cooperative Agreement W911NF-19-2-0026 and contract W911NF-19-D-0001 from the Army Research Office. The content of the information does not necessarily reflect the position or the policy of the Government and no official endorsement should be inferred.

## Declaration of Competing Interests

The authors declare no competing interests.

## Acknowledgments

The authors thank Jan Martin and Frederik Bernd Lund at the Institute of Radiology, University Hospital Erlangen, Friedrich-Alexander-Universität Erlangen-Nürnberg (FAU), Erlangen, Germany for providing software code for the gradient tensor waveforms. We would also like to thank Mario Mendoza for MRI data collection and participant recruitment assistance.

## Supplementary Material

**Figure S1.**
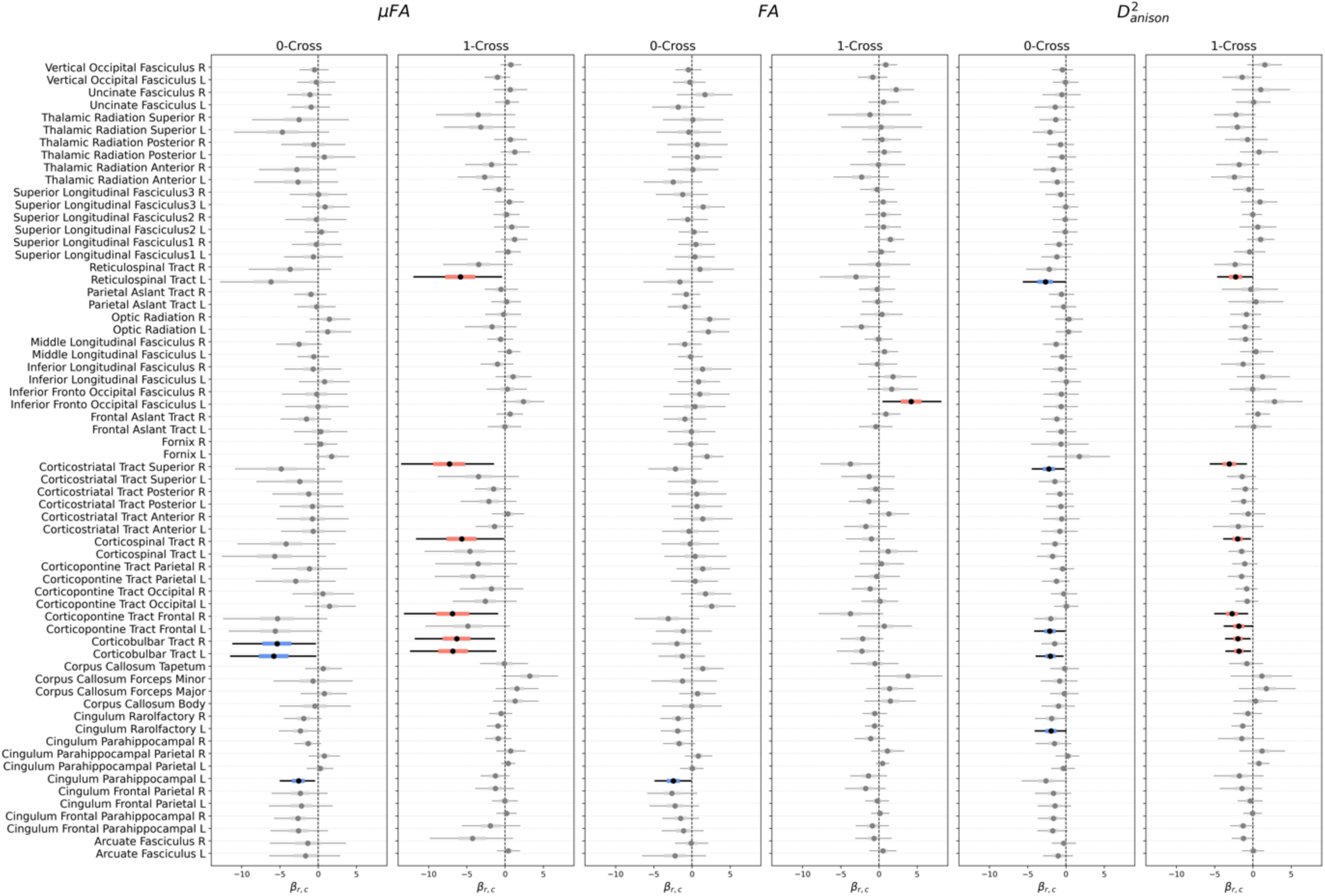
Region-cross model posteriors for MDM tensor shape parameters. Credible β_r,c_ posteriors (94% HDI does not contain 0) for 0-Cross subregions are bolded and shaded blue; β_r,c_ posteriors for 1-Cross subregions are bolded and shaded salmon. Positive β_r,c_ indicates that concussion group membership is more likely (greater-log odds) with greater values of that diffusion parameter in that region-cross ROI. In contrast, negative β_r,c_ indicates that concussion group membership is more likely (greater-log odds) with lower values of that parameter in that region-cross ROI.

**Figure S2.**
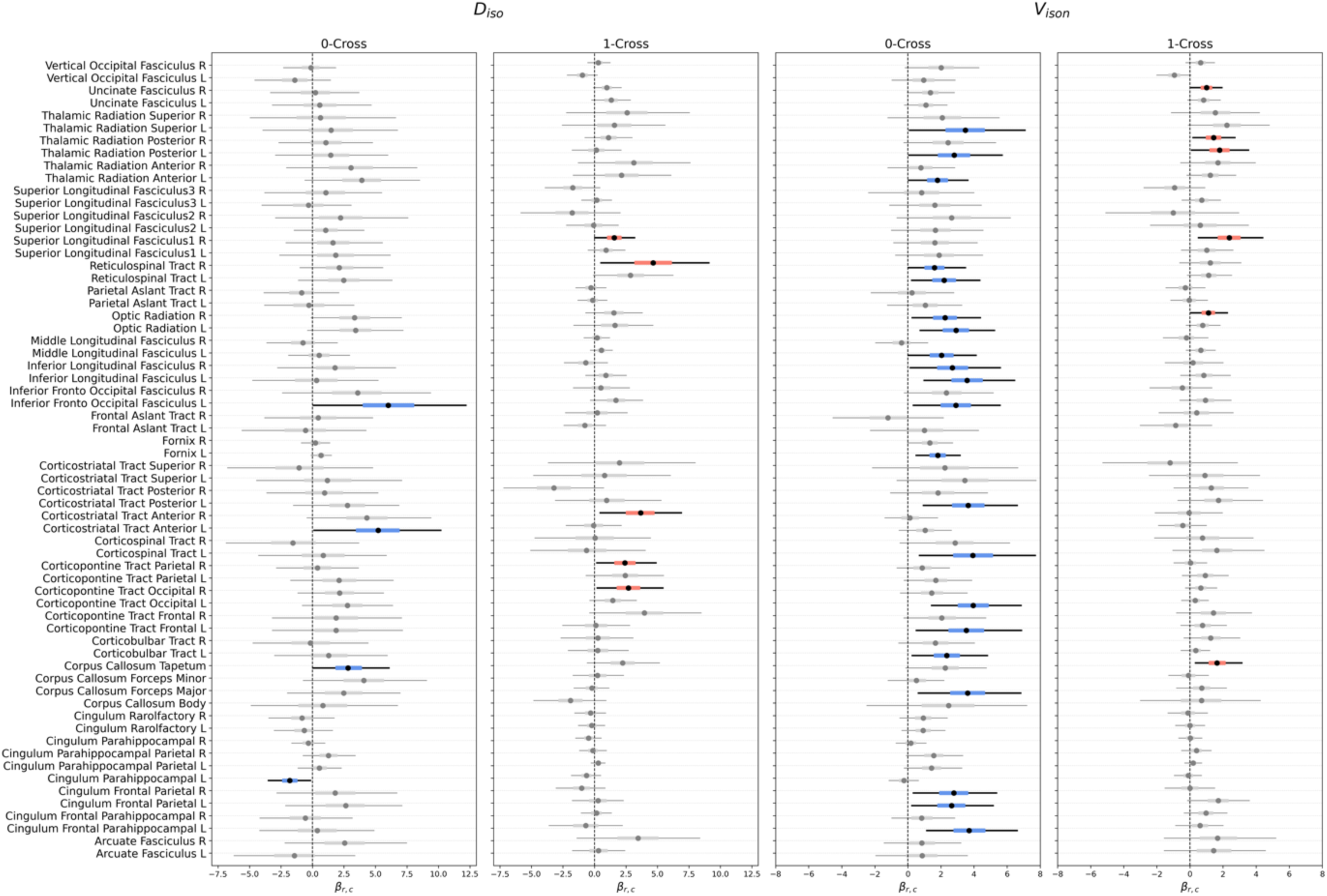
Region-cross model posteriors for MDM tensor size parameters. Credible β_r,c_ posteriors (94% HDI does not contain zero) for 0-Cross subregions are bolded and shaded blue; β_r,c_ posteriors for 1-Cross subregions are bolded and shaded salmon. Positive β_r,c_ indicates that concussion group membership is more likely (greater-log odds) with greater values of that diffusion parameter in that region-cross ROI. In contrast, negative β_r,c_ indicates that concussion group membership is more likely (greater-log odds) with lower values of that parameter in that region-cross ROI.

**Table S1.**
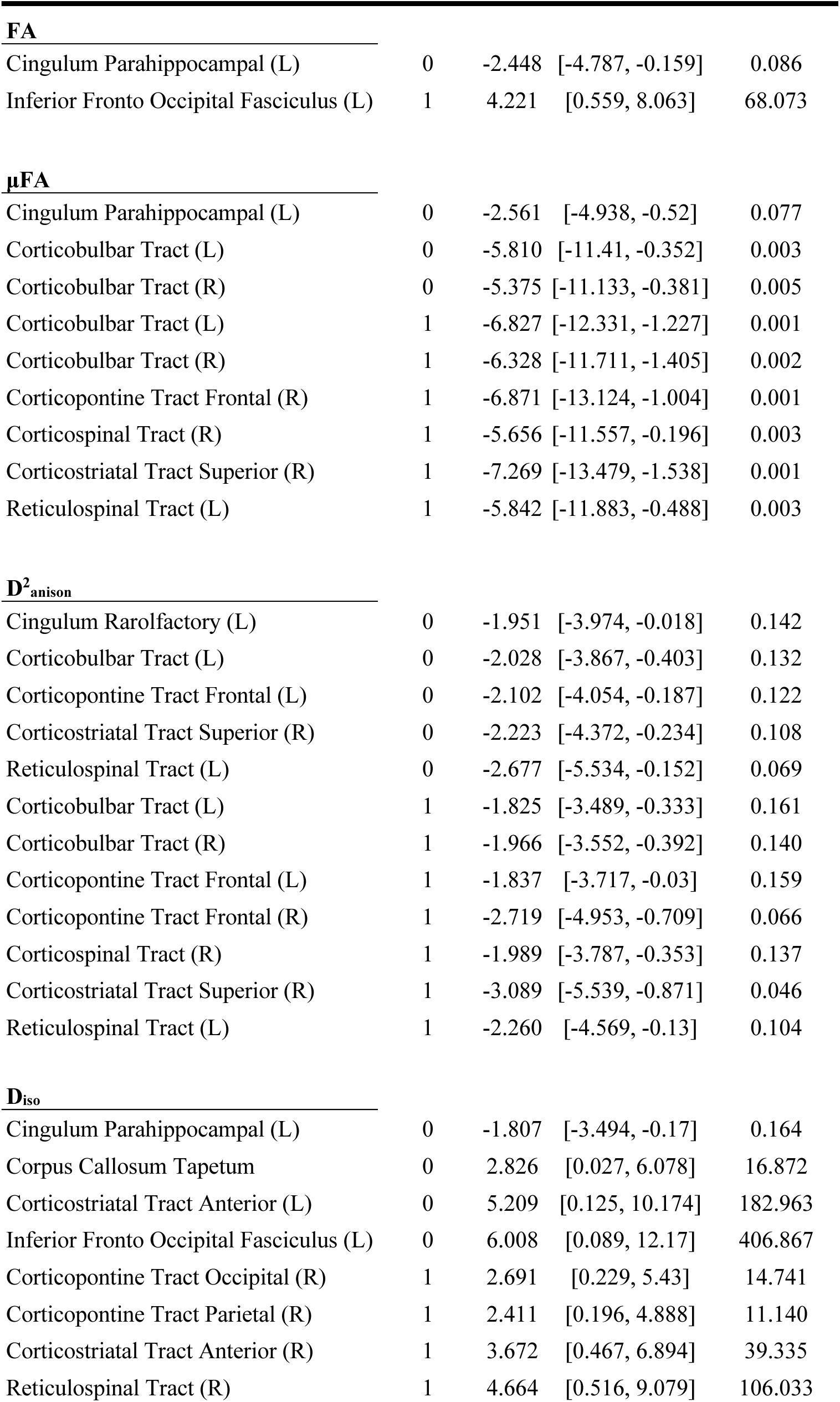
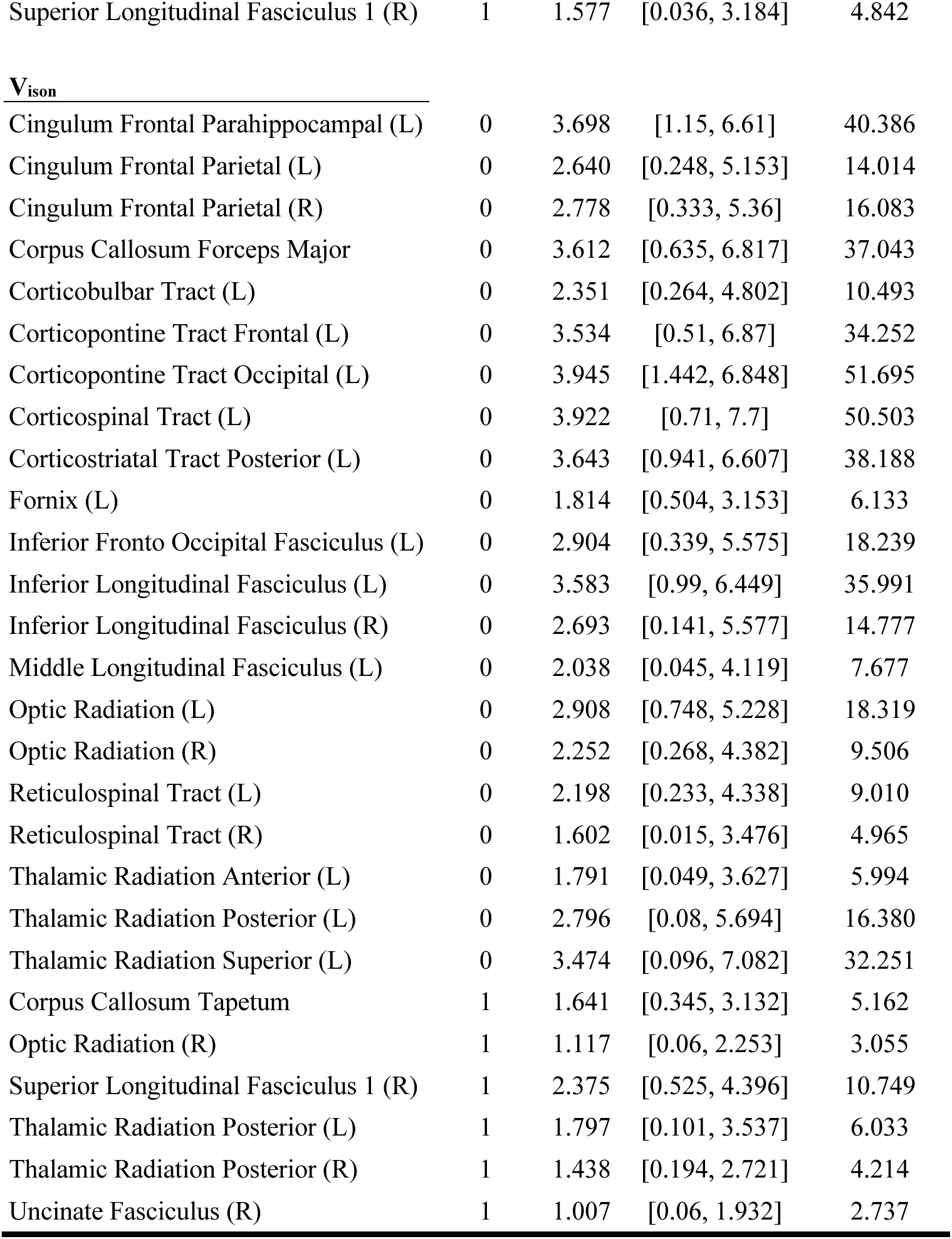
Region-cross model: Credible logistic regression diffusion β posteriors.

| Region Name | Cross ID | $\beta_{r,c}$ | HDI | Odds Ratio |
| --- | --- | --- | --- | --- |
| **FA** |  |  |  |  |
| Cingulum Parahippocampal (L) | 0 | -2.448 | [-4.787, -0.159] | 0.086 |
| Inferior Fronto Occipital Fasciculus (L) | 1 | 4.221 | [0.559, 8.063] | 68.073 |
| **μFA** |  |  |  |  |
| Cingulum Parahippocampal (L) | 0 | -2.561 | [-4.938, -0.52] | 0.077 |
| Corticobulbar Tract (L) | 0 | -5.810 | [-11.41, -0.352] | 0.003 |
| Corticobulbar Tract (R) | 0 | -5.375 | [-11.133, -0.381] | 0.005 |
| Corticobulbar Tract (L) | 1 | -6.827 | [-12.331, -1.227] | 0.001 |
| Corticobulbar Tract (R) | 1 | -6.328 | [-11.711, -1.405] | 0.002 |
| Corticopontine Tract Frontal (R) | 1 | -6.871 | [-13.124, -1.004] | 0.001 |
| Corticospinal Tract (R) | 1 | -5.656 | [-11.557, -0.196] | 0.003 |
| Corticostriatal Tract Superior (R) | 1 | -7.269 | [-13.479, -1.538] | 0.001 |
| Reticulospinal Tract (L) | 1 | -5.842 | [-11.883, -0.488] | 0.003 |
| **$D^2_{anison}$** |  |  |  |  |
| Cingulum Rarolfactory (L) | 0 | -1.951 | [-3.974, -0.018] | 0.142 |
| Corticobulbar Tract (L) | 0 | -2.028 | [-3.867, -0.403] | 0.132 |
| Corticopontine Tract Frontal (L) | 0 | -2.102 | [-4.054, -0.187] | 0.122 |
| Corticostriatal Tract Superior (R) | 0 | -2.223 | [-4.372, -0.234] | 0.108 |
| Reticulospinal Tract (L) | 0 | -2.677 | [-5.534, -0.152] | 0.069 |
| Corticobulbar Tract (L) | 1 | -1.825 | [-3.489, -0.333] | 0.161 |
| Corticobulbar Tract (R) | 1 | -1.966 | [-3.552, -0.392] | 0.140 |
| Corticopontine Tract Frontal (L) | 1 | -1.837 | [-3.717, -0.03] | 0.159 |
| Corticopontine Tract Frontal (R) | 1 | -2.719 | [-4.953, -0.709] | 0.066 |
| Corticospinal Tract (R) | 1 | -1.989 | [-3.787, -0.353] | 0.137 |
| Corticostriatal Tract Superior (R) | 1 | -3.089 | [-5.539, -0.871] | 0.046 |
| Reticulospinal Tract (L) | 1 | -2.260 | [-4.569, -0.13] | 0.104 |
| **$D_{iso}$** |  |  |  |  |
| Cingulum Parahippocampal (L) | 0 | -1.807 | [-3.494, -0.17] | 0.164 |
| Corpus Callosum Tapetum | 0 | 2.826 | [0.027, 6.078] | 16.872 |
| Corticostriatal Tract Anterior (L) | 0 | 5.209 | [0.125, 10.174] | 182.963 |
| Inferior Fronto Occipital Fasciculus (L) | 0 | 6.008 | [0.089, 12.17] | 406.867 |
| Corticopontine Tract Occipital (R) | 1 | 2.691 | [0.229, 5.43] | 14.741 |
| Corticopontine Tract Parietal (R) | 1 | 2.411 | [0.196, 4.888] | 11.140 |
| Corticostriatal Tract Anterior (R) | 1 | 3.672 | [0.467, 6.894] | 39.335 |
| Reticulospinal Tract (R) | 1 | 4.664 | [0.516, 9.079] | 106.033 |
| Superior Longitudinal Fasciculus 1 (R) | 1 | 1.577 | [0.036, 3.184] | 4.842 |
| **Vison** |  |  |  |  |
| Cingulum Frontal Parahippocampal (L) | 0 | 3.698 | [1.15, 6.61] | 40.386 |
| Cingulum Frontal Parietal (L) | 0 | 2.640 | [0.248, 5.153] | 14.014 |
| Cingulum Frontal Parietal (R) | 0 | 2.778 | [0.333, 5.36] | 16.083 |
| Corpus Callosum Forceps Major | 0 | 3.612 | [0.635, 6.817] | 37.043 |
| Corticobulbar Tract (L) | 0 | 2.351 | [0.264, 4.802] | 10.493 |
| Corticopontine Tract Frontal (L) | 0 | 3.534 | [0.51, 6.87] | 34.252 |
| Corticopontine Tract Occipital (L) | 0 | 3.945 | [1.442, 6.848] | 51.695 |
| Corticospinal Tract (L) | 0 | 3.922 | [0.71, 7.7] | 50.503 |
| Corticostriatal Tract Posterior (L) | 0 | 3.643 | [0.941, 6.607] | 38.188 |
| Fornix (L) | 0 | 1.814 | [0.504, 3.153] | 6.133 |
| Inferior Fronto Occipital Fasciculus (L) | 0 | 2.904 | [0.339, 5.575] | 18.239 |
| Inferior Longitudinal Fasciculus (L) | 0 | 3.583 | [0.99, 6.449] | 35.991 |
| Inferior Longitudinal Fasciculus (R) | 0 | 2.693 | [0.141, 5.577] | 14.777 |
| Middle Longitudinal Fasciculus (L) | 0 | 2.038 | [0.045, 4.119] | 7.677 |
| Optic Radiation (L) | 0 | 2.908 | [0.748, 5.228] | 18.319 |
| Optic Radiation (R) | 0 | 2.252 | [0.268, 4.382] | 9.506 |
| Reticulospinal Tract (L) | 0 | 2.198 | [0.233, 4.338] | 9.010 |
| Reticulospinal Tract (R) | 0 | 1.602 | [0.015, 3.476] | 4.965 |
| Thalamic Radiation Anterior (L) | 0 | 1.791 | [0.049, 3.627] | 5.994 |
| Thalamic Radiation Posterior (L) | 0 | 2.796 | [0.08, 5.694] | 16.380 |
| Thalamic Radiation Superior (L) | 0 | 3.474 | [0.096, 7.082] | 32.251 |
| Corpus Callosum Tapetum | 1 | 1.641 | [0.345, 3.132] | 5.162 |
| Optic Radiation (R) | 1 | 1.117 | [0.06, 2.253] | 3.055 |
| Superior Longitudinal Fasciculus 1 (R) | 1 | 2.375 | [0.525, 4.396] | 10.749 |
| Thalamic Radiation Posterior (L) | 1 | 1.797 | [0.101, 3.537] | 6.033 |
| Thalamic Radiation Posterior (R) | 1 | 1.438 | [0.194, 2.721] | 4.214 |
| Uncinate Fasciculus (R) | 1 | 1.007 | [0.06, 1.932] | 2.737 |

## Notes

### Competing Interest Statement

The authors have declared no competing interest.

### Author Declarations

The Institutional Review Board of the University of California, Santa Barbara gave ethical approval for this work.

